# Interpretational fallacies in sibling comparison designs with social exposures: A case study of childhood income and mental disorders

**DOI:** 10.1101/2023.06.23.23291796

**Authors:** Linda Ejlskov, Buket Öztürk Esen, Christian Hakulinen, Nanna Weye, Tomas Formanek, John J. McGrath, Carsten Bøcker Pedersen, Oleguer Plana-Ripoll

**Author notes:** **Corresponding author:** Oleguer Plana-Ripoll.

## Abstract

Sibling comparison designs are increasingly used to address unmeasured familial confounding in observational studies. We propose that three key interpretational fallacies – sibling characteristics, exposure correlation and non-shared confounding, and unmet life course model assumptions – can mislead causal conclusions. We demonstrate these fallacies by investigating childhood family income and mental health.

A nationwide Danish cohort of individuals born between 1986 and 1996 (n = 643,814; 404,179 siblings) was followed-up from age 15 until onset of severe mental disorders. Population-wide and within-sibling adjusted hazard ratios (aHR) between childhood family income and offspring mental disorders were estimated, supplemented by descriptive statistics and pseudo-sibling analyses.

A $15,000 increase in family income at age 14 was associated with a reduced rate of severe mental disorders (aHR = 0.78; 95% CI: 0.76-0.81), with comparable estimates across measurement ages 1-14 (range: 0.67-0.82). Null results were observed in both a pseudo-sibling cohort of unrelated individuals with the same income differences as the true sibling cohort (aHR = 0.93; 95% CI: 0.85-1.01) and the true sibling cohort (aHR = 1.02; 95% CI: 0.94-1.11). Siblings were typically born three years apart, with an average monthly income difference of $496 at age 14 (IQR;$150-$641).

This study advocates for cautious causal interpretation of null results in sibling comparison studies because (1) it may not capture meaningful differences in family income across siblings due to minor income fluctuations; (2) the pseudo-sibling cohort showed evidence of unmeasured non-shared confounding; (3) sibling comparison designs test a critical periods life course model, but the results favour an accumulating/vulnerable model. We present guidelines and R syntax to assess these interpretational fallacies.

## Background

Strong observational evidence has consistently demonstrated associations between lower family income and an increased risk of various adverse outcomes^1,2^, including mental disorders.^3–5^ However, the causal nature of this association remains the subject of ongoing debate^6^, partly due to the possibility of unmeasured familial confounding. To address this concern, sibling comparison designs – in which each individual is compared only to their siblings – are increasingly used in observational studies examining social exposures like family income.^7^

A typical approach to interpreting such sibling comparison designs studies involves identifying an association between family income at e.g. age 14 and later mental health outcomes in a general population.^8^ This association is then re-examined within sibling pairs to account for unmeasured familial confounding. The interpretation is that if the association is causal, it should persist within siblings who share the same family environment, as shared genetic and environmental factors are controlled for in these comparisons.^9^ While this design has the appeal of controlling for stable confounding variables across siblings, recent studies have raised concerns about its interpretive validity.^7,9–11^ Specifically, there is a risk of biased estimates when the exposure is highly correlated between siblings.^12,13^ In addition to these well-documented limitations, we propose that three key interpretational fallacies inherent to sibling comparison designs can lead to misleading conclusions about causality, especially in the context of null or attenuated results. The three fallacies – sibling characteristics; exposure correlation and unmeasured non-shared confounding; and unmet life course models assumptions – all follow from the basic premise of the sibling comparison design. Namely, it compares family income measured at the same age for each sibling and estimates whether the difference between these measurements is associated with differences in the risk of having a mental disorder. As illustrated in Figure 1, because they are siblings, it follows that the family income that one sibling experiences at one age is the same family income that the other sibling experiences at another age, typically three years older or younger.^14^

**Figure 1.**
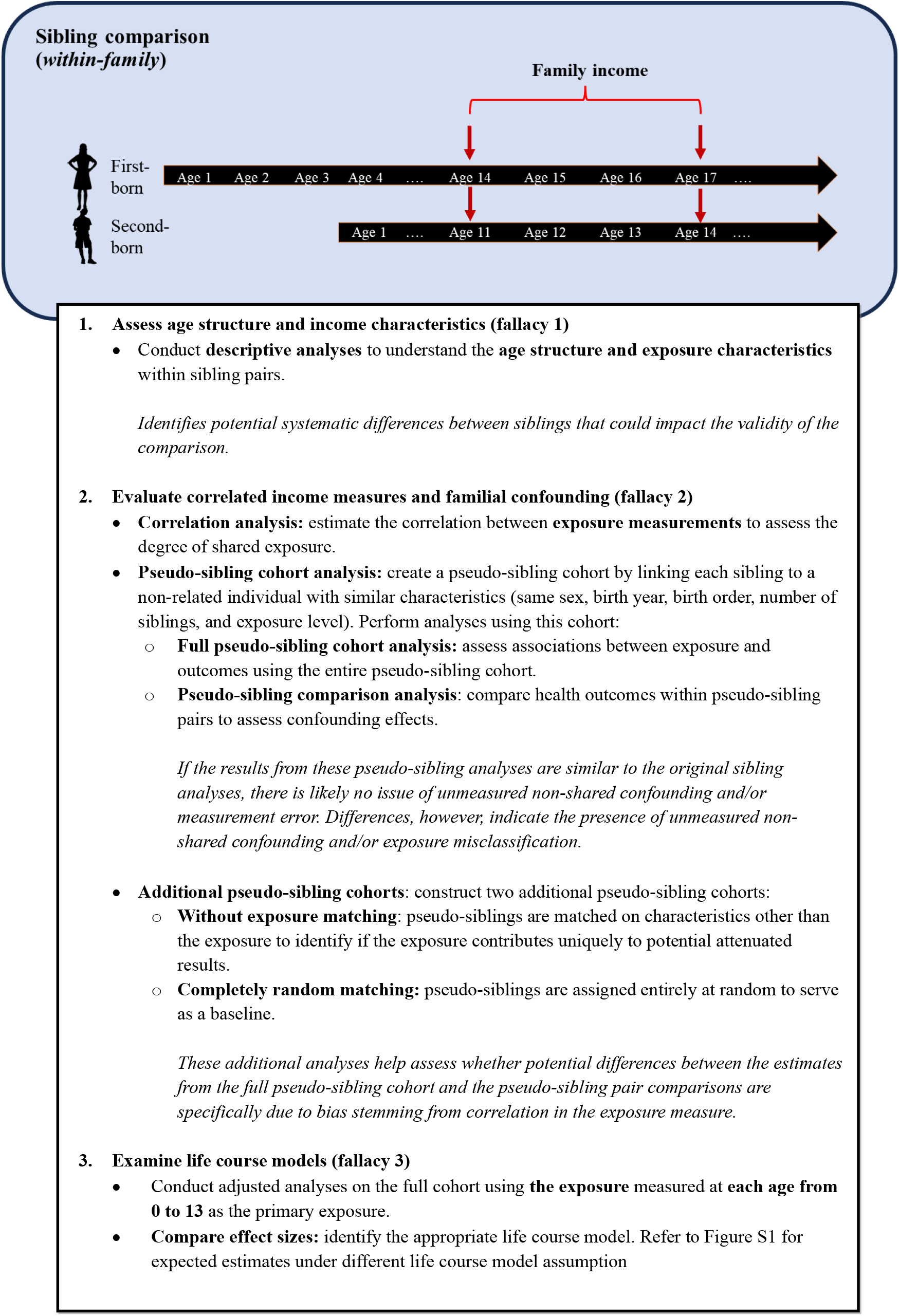
Guide to assess interpretational fallacies in sibling comparison studies.

The first fallacy arises from the assumption that sibling comparison designs can capture differential exposure to family income across siblings. Thus, this approach assumes that income differences between distinct points in each sibling’s life course can lead to meaningful differences in their lived experiences, such as differential changes in parenting behavior or environmental conditions that affect each sibling’s risk of developing an adverse health outcome. Thus, characteristics of the sibling pairs themselves, such as age gaps, income characteristics, and income differences can greatly inform the interpretation of the results of sibling comparison studies depending on the typical age gap between siblings, whether the income is already sufficient to cover basic needs, the difference in income between the two timepoints, etc.

The second fallacy arises from the assumption that a high correlation between siblings’ family income measurements does not attenuate results due to either non-shared unmeasured confounding or exposure misclassification. Previous research has shown that, as the correlation between exposure measurements increases, the potential for non-shared unmeasured confounding to distort the results in sibling comparison design also increases.^12,15^ Additionally, exposure misclassification attenuates associations between the exposure and outcome more in sibling comparison designs than unpaired analyses, and this attenuation in sibling comparison designs intensifies as the correlation in exposure between siblings increases.^13^

The third fallacy arises as both siblings live within the same family income environment at the same time just at different ages, then sibling comparison designs implicitly assume a critical period of vulnerability to income-related exposures and risk of later adverse health life course model, in which family income at a specific age (e.g., age 14) has a disproportionate effect on the risk of mental health compared to another age (e.g., age 11). However, if this assumption is not met, null results from a sibling comparison design may simply reflect that experiencing a particular income environment at e.g. age 14 compared to e.g. age 11 does not capture the complex ways in which family income influences mental health.

This paper empirically assesses each of these interpretational fallacies in sibling comparison designs as illustrated in Figure 1. Using data from a nationwide Danish cohort, we investigate the association between childhood family income and the subsequent risk of severe mental disorders in the offspring. Through triangulation of descriptive statistics, pseudo-sibling cohorts, and traditional cohort analyses, we aim to show how not considering the fallacies described above can lead to misleading causal interpretations of the results. Based on the findings we will enable researchers to better interpret estimates from sibling comparison studies in the future.

## Methods

The protocol for this study was pre-registered before any analyses were commenced.^16^ We designed a population-based cohort study including all individuals who were: i) born in Denmark between January 1^st^, 1986 and December 31^st^, 1996, ii) alive and residing in Denmark on 15^th^ birthday, and iii) possible to link to both parents to enable identification of full siblings. The Danish Civil Registration System^17,18^ maintains continuously updated information such as sex, date of birth, emigration, and death of all residents. Everyone living in Denmark is given a unique personal identification number enabling the linkage of relevant information across the Danish registries.^17,18^ We identified the first diagnosis of severe mental disorder (schizophrenia-spectrum disorder and bipolar disorder) after 15^th^ birthday using the Psychiatric Central Research Register (see appendix page 2 for the specific diagnostic codes following Pedersen et al.^19^). The study cohort was followed from 15^th^ birthday until they were diagnosed with a severe mental disorder, emigrated, died or end of follow-up on December 31^st^, 2018, with the longest possible follow-up being 18 years (i.e., followed until age 33 years).

Family income was measured using information on equated disposable household income obtained from the Income Statistics Register.^20^ This measure subtracts expenses such as taxes and interest expenses from taxable income while also correcting for the difference in family size using the OECD modified scale.^21^ Family income was measured the year before the start of follow-up (age 14) for each cohort member in the main analyses and at each year during childhood (0-13 years) for additional analyses. The measure was inflation-adjusted and represented in US dollar ($) values.^22^ Due to some very extreme income values, we chose to exclude individuals in the lowest and highest percentiles (see appendix page 3 and Figure S1 for more detailed information). The final study cohort consisted of 643,814 individuals (98% of the eligible study cohort). Information on sociodemographic factors (birth order, origin, parental education, urbanicity), parental history of psychiatric disorders, and markers of family function and dysfunction (parental incarceration, family cohabitation status) were identified through population registers and based on information until the individual’s 15^th^ birthday (see appendix page 4 for more information on covariates).

## Statistical analyses

The association between family income at age 14 and subsequent risk of developing severe mental disorders in the offspring was estimated through adjusted hazard ratios (aHRs) using Cox proportional hazards models with age as the underlying time scale. We fitted a model in the original cohort adjusting for sex, birth year, birth order, origin, urbanicity, family cohabitation, parental psychiatric disorders, and parental incarceration. We explored potential nuances in the association between disposable family income and the development of severe mental disorders by re-running the model with family income divided in quintiles, divided in deciles, and as the mean income over the whole early life course (0-14 years). Second, we performed a sibling comparison design by fitting a stratified Cox regression model (stratifying on family, including only full siblings) adjusting for the same variables allowing for a comparison of changes in family income and subsequent risk of developing severe mental disorders solely between siblings.^23^

### Additional analyses to assess interpretational fallacies

An overview of the analyses to assess each of the three fallacies is provided in Figure 1.

### Fallacy 1

To assess the first interpretational fallacy, we conducted descriptive analyses to understand the age structure and income characteristics within sibling pairs.

### Fallacy 2

For the second fallacy, which concerns the possible attenuation effect of exposure misclassification and unmeasured non-shared confounding, we conducted two key analyses. First, we estimated the correlation in family income between siblings at age 14 to assess the extent of shared exposure. Second, we wanted to investigate whether the results from the sibling comparison design would be affected by unmeasured non-shared confounding between siblings and/or exposure misclassification. Thus, we created a pseudo-sibling cohort by linking each sibling in the original cohort to a non-related individual. For each pair of siblings, we randomly kept one of the siblings and replaced the other sibling with a random non-related individual who had the same characteristics as the actual sibling in terms of sex, birth year, birth order, number of siblings, and family income (pseudo-sibling cohort γ). In this simulated cohort, we performed both population-wide pseudo-sibling cohort and pseudo-sibling comparison analyses, mirroring the original cohort analyses. Given that sibling-pairs in the pseudo-sibling cohort were created at random, similar estimates from the population-wide pseudo-sibling cohort and pseudo-sibling comparison analysis would indicate no issues of unmeasured non-shared confounding and/or exposure misclassification in the main analysis. In contrast, different estimates between the two would indicate a general problem of unmeasured non-shared confounding or exposure misclassification.^15,23^ We also created two additional pseudo-sibling cohorts for further comparison: one that disregarded income while matching pseudo-siblings (pseudo-sibling cohort β), and another where pseudo-siblings were assigned completely at random (pseudo-sibling cohort α). These analyses helped to clarify whether potential differences between the estimates from the full pseudo-sibling cohort and the pseudo-sibling pair comparisons were due to bias stemming from correlation in the income measure or were derived from other variables.^24^ Appendix page 5 provides more details on the creation of the pseudo-sibling cohorts and Table S1 presents an overview of the three different cohorts used in the study.

### Third fallacy

To investigate the third interpretational fallacy, we investigated whether the assumption of a critical period during which income disproportionately impacts mental health is plausible. Figure S2 illustrates the expectation of estimates from family income measured at different age points under three life course model assumptions. Thus, we conducted a series of separate adjusted analyses in the full cohort, with family income measured at each age from 0 through 13 as the primary exposure. This allowed us to evaluate whether family income at one specific age-point has a disproportionate effect size compared to income measured at other childhood ages. All data analyses were performed using R version 4.21.^25^

## Results

We followed 643,814 individuals for a total of 7.8 million person-years, with 14,637 (2.3%) developing a severe mental disorder during the follow-up period. Table 1 presents the descriptive characteristics of the entire study cohort. Table 2 shows characteristics of the sibling cohort (i.e., those who had at least one sibling in the cohort) by family income decile grouping. The average age difference between siblings was 3.2 years (sd=1.5) and the average annual disposable family income at age 14 was $30,712 (IQR=$24,310;$35,741). The average difference in income between siblings increased with higher family income, while the prevalence of mental disorders decreased. In total, 84% of the sibling cohort had changes in annual disposable income between siblings below $10,000, and only 1.2% of the sibling cohort had differences larger than $30,000 (80% of these belonging to families in the three highest disposable family income deciles) (Table S4). For more details and distributions of sibling characteristics see Appendix, Figures S5-S9, and Tables S2-S4.

**Table 1.** Sociodemographic characteristics of the study cohort (n = 643 814)

| Variable | n | Mental disorder<br>diagnosis<br>n(%) | Disposable family income (\$)<br>median (IQR) |
| --- | --- | --- | --- |
| **Sex** |  |  |  |
| Female | 313,84 | 7,078 (2.2) | 30,310 (24,211 : 37,097) |
| Male | 329,97 | 7,559 (2.2) | 30,432 (24,362 : 37,205) |
| **Number of children** |  |  |  |
| One child | 239,64 | 5,721 (2.3) | 31,594 (25,248 : 38,730) |
| Two children | 311,26 | 6,732 (2.1) | 30,638 (24,837 : 37,049) |
| Three children | 81,327 | 1,891 (2.3) | 27,147 (21,588 : 33,299) |
| Four children or more | 11,588 | 293 (2.5) | 21,781 (17,242 : 27,227) |
| **Birthorder** |  |  |  |
| First-born | 425,19 | 10,157 (2.3) | 30,381 (24,367 : 37,172) |
| Second-born | 185,56 | 3,820 (2.0) | 30,576 (24,466 : 37,291) |
| Third-born | 29,927 | 590 (1.9) | 29,461 (22,807 : 36,506) |
| Fourth-born or higher | 3,134 | 70 (2.2) | 24,535 (19,395 : 31,177) |
| **Parental psychiatric**<br>**disorder history** |  |  |  |
| No | 583,58 | 11,855 (2.0) | 30,812 (24,819 : 37,557) |
| Yes | 60,237 | 2,782 (4.4) | 25,794 (20,410 : 32,365) |
| **Parental incarceration** |  |  |  |
| No | 613,04 | 13,309 (2.1) | 30,749 (24,772 : 37,488) |
| Yes | 30,779 | 1,328 (4.1) | 22,578 (18,081 : 28,012) |
| **Country of origin** |  |  |  |
| Parents born in Denmark | 560 | 12,367 (2.2) | 31,008 (25,183 : 37,663) |
| Parent/s born abroad | 82,864 | 2,251 (2.6) | 25,110 (19,386 : 32,526) |
| Missing | 952 | 19 (2.0) | 26,356 (20,662 : 33,143) |
| **Family cohabitation status** |  |  |  |
| Cohabiting/married | 506,25 | 9,663 (1.9) | 32,122 (26,400 : 38,712) |
| Single | 129,29 | 4,362 (3.3) | 24,611 (20,599 : 29,400) |
| Single living alone | 7,485 | 579 (7.2) | 2,379 (1,096 : 3,414) |
| Missing | 789 | 33 (4.0) | 20,097 (12,017 : 31,242) |
| **Parental educational level** |  |  |  |
| Low | 46,191 | 1,606 (3.4) | 23,523 (18,999 : 28,692) |
| Middle | 342,37 | 7,334 (2.1) | 29,488 (24,065 : 35,276) |
| High | 213,53 | 4,321 (2.0) | 34,826 (28,564 : 42,472) |
| Missing | 41,726 | 1,376 (3.2) | 24,346 (18,922 : 30,972) |

**Table 2.** Family characteristics for 404 179 individuals included in the sibling cohort by disposable family income deciles.

| | n | Severe mental disorder<br>n(%) | Mean number of children (sd) | Mean age difference (sd) | Family income (\$) difference between measurements | |
| --- | --- | --- | --- | --- | --- | --- |
|  |  |  |  |  | Mean | Median (IQR) |
| Overall | 404,179 | 8,916 (2.2) | 2.3 (0.5) | 3.2 (1.5) | 5,957 | 3,945 (1,807 : 7,701) |
| **Disposable family** |  |  |  |  |  |  |
| Lowest decile | 42,068 | 1,648 (3.9) | 2.5 (0.7) | 2.9 (1.5) | 6,296 | 3,561 (1,662 : 8,259) |
| 2 | 42,507 | 1,355 (3.2) | 2.4 (0.6) | 3.1 (1.5) | 4,923 | 3,481 (1,741 : 6,235) |
| 3 | 43,029 | 1,117 (2.6) | 2.3 (0.5) | 3.2 (1.5) | 4,856 | 3,540 (1,695 : 6,372) |
| 4 | 43,405 | 927 (2.1) | 2.3 (0.5) | 3.2 (1.4) | 4,826 | 3,494 (1,634 : 6,476) |
| 5 | 42,810 | 857 (2.0) | 2.3 (0.5) | 3.3 (1.4) | 5,005 | 3,598 (1,682 : 6,721) |
| 6 | 42,139 | 758 (1.8) | 2.2 (0.4) | 3.3 (1.4) | 5,179 | 3,680 (1,700 : 7,061) |
| 7 | 40,792 | 667 (1.6) | 2.2 (0.4) | 3.4 (1.5) | 5,439 | 3,844 (1,724 : 7,363) |
| 8 | 38,820 | 605 (1.6) | 2.2 (0.4) | 3.4 (1.5) | 6,174 | 4,267 (1,860 : 8,359) |
| 9 | 36,072 | 515 (1.4) | 2.1 (0.4) | 3.4 (1.5) | 7,355 | 4,968 (2,195 : 9,711) |
| Highest decile | 32,537 | 467 (1.4) | 2.1 (0.4) | 3.3 (1.5) | 10,932 | 7,572 (3,256 : 15,125) |

Note: To be included in the sibling cohort, one had at least one sibling in the full population-wide cohort

Figure 2 presents the results for the association between annual disposable family income and subsequent rate of developing severe mental disorders based on the four different cohorts. Based on the original cohort, for each $15,000 increase in disposable family income measured at age 14, the rate of developing a severe mental disorder was reduced by 22% in the population-wide analysis (aHR=0.78; 0.76-0.81). When investigating the same association solely within siblings exposed to different levels of disposable family income at age 14, the association was entirely attenuated (aHR=1.02; 0.94-1.11). Results were robust to different family income categorisations and when only using those from the sibling cohort (Table S2, Figures S3-S4).

**Figure 2.**
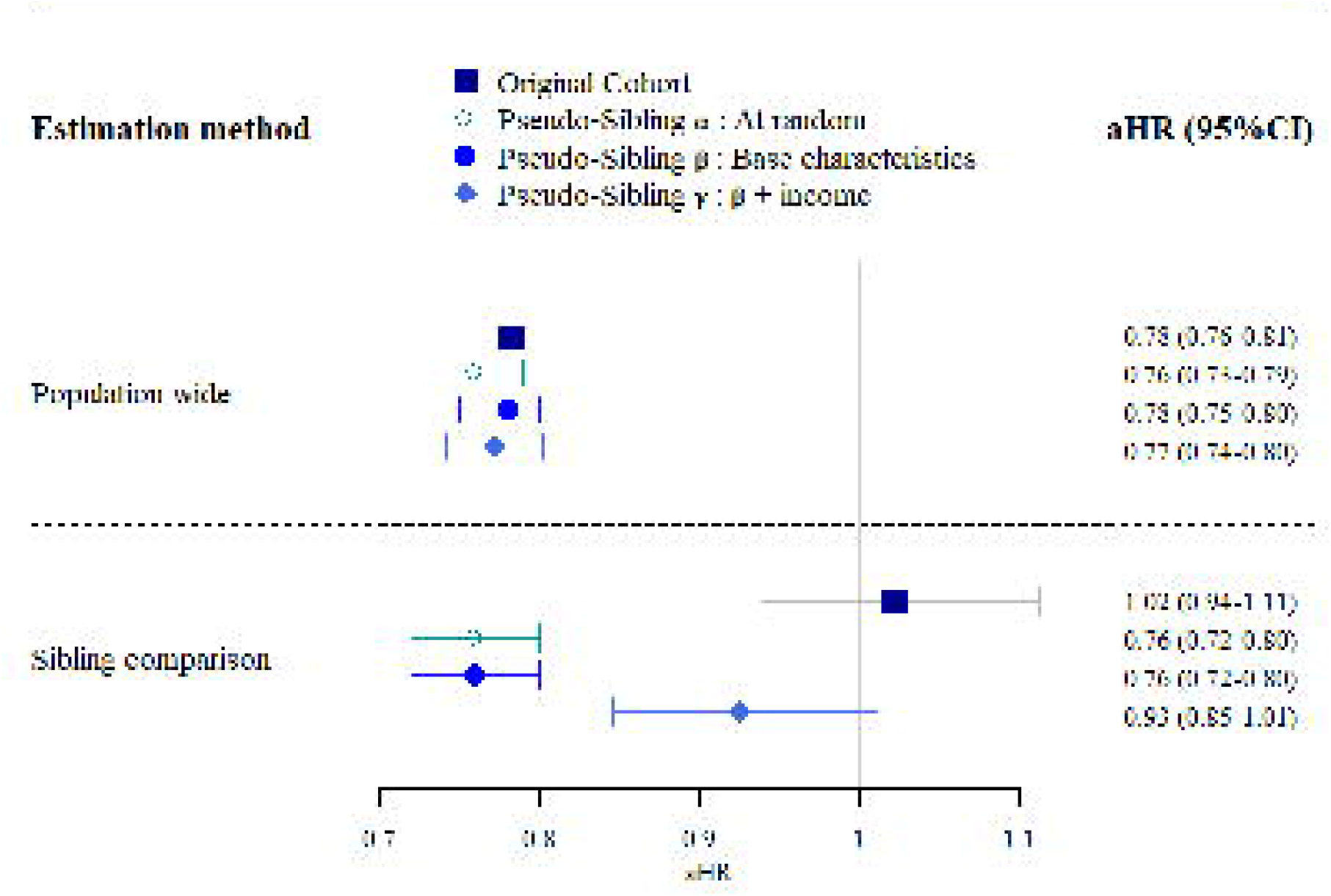
Adjusted hazard ratios (aHR) and 95% confidence intervals (CI) between disposable family income measured at age 14 (in units of $15,000) and subsequent severe mental disorders. The estimates from full cohort analyses (model 1) and sibling comparison designs for the original cohort (models 1 and 2) and the simulated pseudo-sibling cohorts (models 3-8). Note: Both the *adjusted model* and the *sibling stratified model* are adjusted for sex, birth order, birth year, origin, parental incarceration, parental psychiatric history, urbanicity and family cohabitation status. The *sibling stratified model* has a baseline hazard stratified by family ID.

In the original cohort, the correlation in family income at age 14 years between siblings in each family was 0.67. Within both pseudo-siblings matched at random (α) or based on the same base characteristics (β), there was no correlation in family income between pseudo-siblings (0.01 and 0.04) and similar estimates using the population-wide and sibling comparison design. Pseudo-sibling cohort γ additionally included income in the pseudo-sibling matching process: the correlation in family income between these pseudo-siblings was 0.65. The population-wide estimate was similar to the other cohorts (aHR = 0.77; 0.74-0.80). However, when using the sibling comparison design, the association was heavily attenuated (aHR = 0.93; 95%CI: 0.85-1.01), although these pseudo-siblings did not belong to the same families. Figure 3 presents the results for the population-wide association between disposable family income measured across ages 1– 14 years and the subsequent risk of developing severe mental disorders based on the original cohort. The associations varied slightly according to the ages at which family income was measured, with aHR ranging from 0.67 to 0.82, and with an aHR for the cumulated family income across the entire childhood period (ages 1 through age 14) of 0.62 (95%CI: 0.60-0.65).

**Figure 3.**
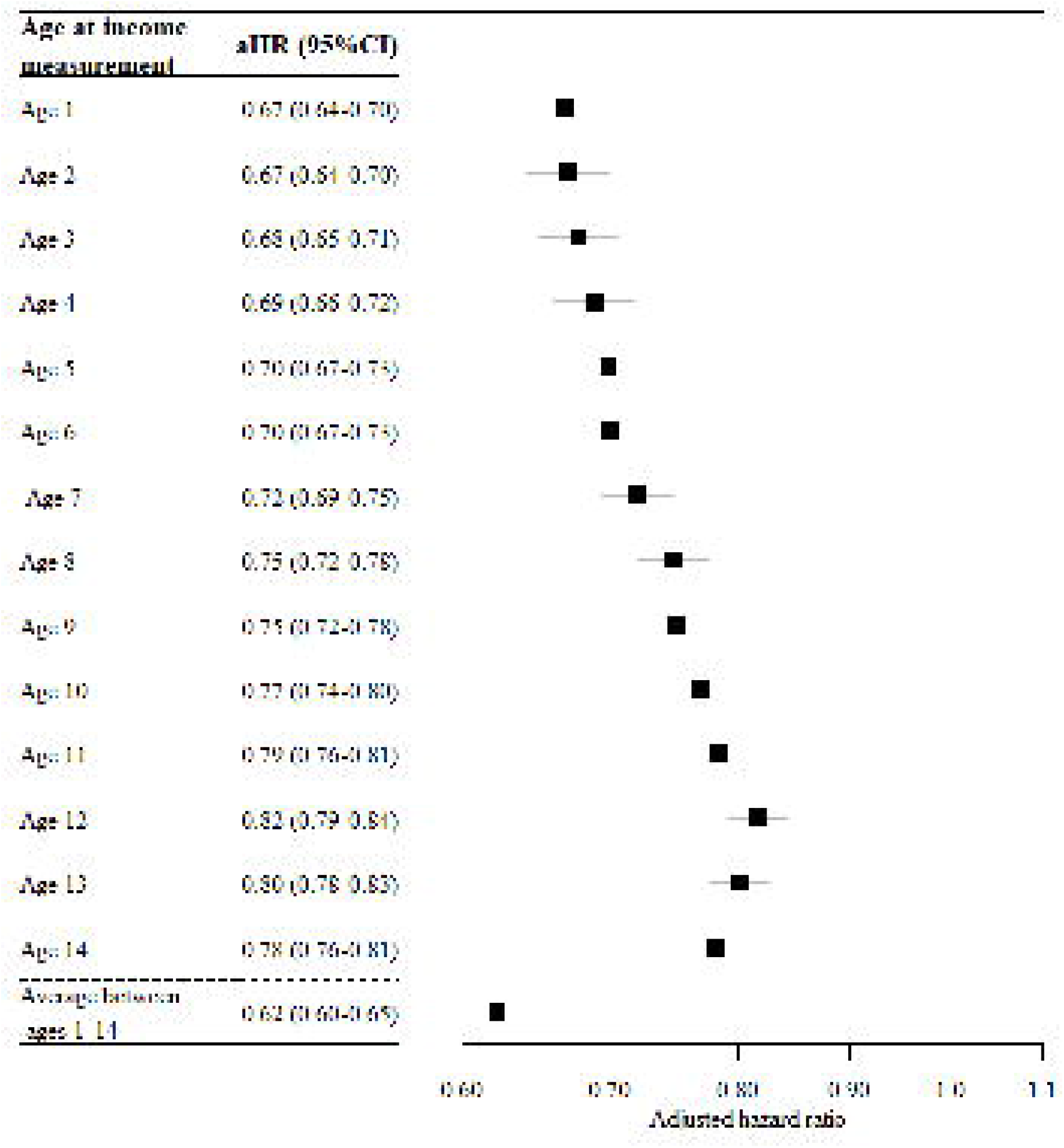
Adjusted hazard ratios (aHR) and 95% confidence intervals (CI) between disposable family income measured across ages 1–14 years (in units of $15,000) and subsequent severe mental disorders. The estimates were obtained from full cohort analyses in the original population (model 1). Note: All models were adjusted for sex, birth order, birth year, origin, parental incarceration, parental psychiatric history, urbanicity and family cohabitation status.

## Discussion

Our study identified and assessed three interpretational fallacies inherent in sibling comparison designs investigating social exposures which, if not assessed, can lead to misleading causal interpretations We found several novel findings important for the interpretation of results from such designs. We demonstrated that while sibling comparison designs are powerful in controlling for shared familial factors, results from these studies are vulnerable to: sibling characteristics; exposure correlation and unmeasured non-shared confounding; and unmet life course models assumptions.

First, we showed that the sibling comparison design compared the risk of developing severe mental disorders for sibling pairs with relatively small differences in age and family income at age 14. The changes in income between the two measurement points in each sibling’s life course were minor, most siblings had a difference in annual family income below $10,000 from one age to the other. Engzell and Hällsten^7^ argue that such minor income fluctuations can be seen as transitory and as a form of measurement error and thus not reflecting true changes in income. Additionally, most sibling pairs resided in a family environment with a baseline income above the Danish Financial Supervisory Authority’s recommendations for adequate living standards.^26^ This raises the question of whether the differences in family income between sibling pairs at age 14 were sufficient to influence differential parental practices or resource allocation toward siblings, subsequently differentially affecting their risk of developing mental disorders. A more probable scenario may be that differential parenting practices towards siblings are based on familial factors unrelated to minor income fluctuations.^27^ For example, a Danish study showed that complex sibling characteristics (age difference, birth order, number of younger siblings, number of older siblings) influenced schizophrenia risk, suggesting that siblings are less comparable than indirectly anticipated in sibling comparison studies.^28^

The second key finding was the high correlation between family income measurements which complicates interpretation. First, when most siblings have similar exposures, then siblings that differ in exposure are also more likely to differ on some non-shared factors that might affect the exposure.^15,23^ An interesting observation from our study was that the families with the highest changes in income between measurements and therefore also those weighing the most in the analyses resided in the highest income families. These families also had the lowest rates of mental disorders overall. If there are other unmeasured factors that both caused the siblings to differ in their family income and risk of mental disorder, then confounding would be larger among siblings that differ in exposure than in the general population. Second, even if no unmeasured non-shared confounding exists, potential exposure misclassification may have attenuated results from the sibling comparison design due to the high correlation between income measurements.^12^ The possibility of attenuation of results by unmeasured non-shared confounding and/or measurement error was supported by the results from the simulated pseudo-sibling cohorts. In these cohorts, different non-related individuals were grouped into pseudo sibling-pairs in three different ways; at random, matching on base characteristics and matching on both base characteristics and family income. In the cohort matching pseudo-siblings on both base characteristics and income, the pseudo-sibling model provided heavily attenuated associations in the sibling comparison analysis, although these individuals grouped into pseudo-sibling pairs did not belong to the same families. However, in the additional pseudo-sibling cohorts that did not consider income to create the pseudo-siblings, the estimates from the full cohort and the sibling comparison design were very similar. This finding lends support to the hypothesis that the findings based on sibling comparisons are biased due to unmeasured non-shared confounding and/or measurement error (probably due to the high correlation in exposure between siblings).

The third key finding was the observation that family income measured at each age during childhood (ages 1 through 14) was associated with the development of a severe mental disorder, showing fairly similar associations across the childhood life course. As illustrated in Figure S2, such estimates support the accumulating/vulnerable risk model and not the critical period model, which is in line with the findings from previous studies.^29,30^ Thus, the results indicate that it is not the timing of a specific family income level that is important. Instead, it is the prolonged exposure to income during the childhood period that is associated with the risk of severe mental disorders. The purpose of the sibling comparison design is to examine whether siblings differ in their risk of developing severe mental disorders based on differences in the family disposable income at age 14.^31^ That is, sibling comparisons designs are useful to assess whether experiencing a specific family income at some specific age (as opposed to the age when the other sibling experiences the same family income) affects health. Because no specific family income measurement age has a disproportionate association with severe mental disorders, the sibling comparison design may be limited in its ability to detect any potential true association between family income and later mental disorders, and instead introduces null results by design. Consequently, other types of study designs, e.g. quasi-experimental designs^4^ or comparing adoptees to biologically born children^5^ may be better methodological alternatives to provide evidence of causality.

In conclusion, we recommend caution in interpreting sibling comparison studies as definitive evidence for or against causal relationships between income and health. Our study suggests that incorporating life course considerations and assessing sibling characteristics can improve the interpretability of sibling comparison design. Moreover, the use of pseudo-sibling cohorts can help differentiate whether potential attenuated effects from a sibling comparison design are due to unmeasured non-shared confounding, measurement error or other factors. We provide a practical guide for researchers with accompanying R-syntax to assess the interpretability of future sibling comparison analyses.

## Data Availability

The individual-level data used for this study are not publicly available but can be obtained by application to Statistics Denmark and the Danish Health Data Authority.

## Ethics approval

This study has been approved by the Danish National Board of Health, and the Danish Data Protection Agency. No informed consent is needed for a register-based study based on de-identified data in Denmark.

## Supplementary data

Supplementary data are available

## Author contributions

LE and OP-R designed and conceptualized the study and drafted the first version of the analyses plan. BÖE, CH, TF and CBP revised the analyses plan and approved the final version. LE did the analyses with input from OP-R. NW checked the R-syntax for potential errors. OP-R supervised the study. LE and OP-R prepared the original draft of the manuscript with input from BÖE and JJM. All other authors provided feedback and suggestions for additional analyses, and reviewed and edited the manuscript. All authors approved the final version.

## Funding

This work was supported by a grant from Independent Research Fund Denmark (grant 1030-00085B). Oleguer Plana-Ripoll is also supported by a Lundbeck Foundation Fellowship (R345-2020-1588) and Independent Research Fund Denmark (grant 2066-00009B). Carsten Bøcker Pedersen is supported by the Big Data Centre for Environment and Health funded by the Novo Nordisk Foundation Challenge Programme (grant NNF17OC0027864). Christian Hakulinen was supported by the European Union (ERC, MENTALNET, 101040247).

The funders of the study had no role in developing the methodology, study design, data collection, data analysis, data interpretation, or writing of the report.

## Conflict of interest

None declared

## Notes

### Competing Interest Statement

The authors have declared no competing interest.

### Author Declarations

This study is approved by the Danish Data Protection Agency. No further ethical approval is required for registry-based research in Denmark.cf. LBK nr 1338 af 01/09/2020, sektion 10 Bekendtgoerelse af lov om videnskabsetisk behandling afsundhedsvidenskabelige forskningsprojekter og sundhedsdatavidenskabelige forskningsprojekter [Act no. 1338 of 1 September 2020, section 10 on research ethics for administration of health scientific research projects and health data scientific research projects].

### Summary of Updates

We have reframed the paper to be more general and now provide guidelines to assess three interpretational fallacies in sibling comparison designs

