## Appendix for "Interpretational fallacies in sibling comparison designs with social exposures: A case study of childhood income and mental disorders"

The causal effect of income of mental health revisited. A nationwide Danish study on the use of sibling comparison designs

#### Contents

**ICD codes to identify severe mental disorder** **2**

**Study population - Income misclassification and outliers** **3**

**Covariates** **4**

**Creation of simulated cohorts** **5**

**Figures** **6**

***Figure S7***. Overall (left) and absolute (right) annual disposable family income differences between siblings measured at age 14 with mean, median and interquartile range (IQR) ($n$ = 404,177) 11

#### ICD codes to identify severe mental disorder

| | ICD 10-DCR codes | Equivalent ICD-8 codes |
|---|---|---|
| **Severe mental disorder** | | |
| Schizophrenia and related disorders | F20-F29 | 295.x9, 296.89, 297.x9, 298.29-298.99, 299.04, 299.05, 299.09, 301.83 |
| Bipolar disorder | F30-F31 | 296.19, 296.39, 298.19 |

#### Study population - Income misclassification and outliers

Figure S1 shows the distribution of annual disposable family income measured at age 14 for the entire study cohort (top row) and the 98%-tile (bottom row) in $. Panels A and C shows the family income distribution while panels B and D shows the amount of family assets for the various disposable family incomes.

As panel A illustrates, there are some very extreme annual taxable income values reported. As we are including family income as a continuous variable in the analyses, this issue is especially pertinent in this study. Especially for the very low/negative reported taxable income, there is an issue of misclassification. Families reporting negative values are often citizens who have stocks, companies or other assets that makes it possible for them to subtract from other incomes and thus report less taxable income. Panel B shows the relationship between the amount of family disposable income and corresponding amount of family assets. Panel B illustrates that as the disposable family income nears zero, the amount of family assets starts to rise drastically with lower reported taxable family incomes, demonstrating the issue that taxable income may not be a good measure to classify the socioeconomic position of these individuals. To ensure that extreme positive or negative values do not weigh too much in the analyses, the lowest and highest 1% of the family income values are removed from the analyses reported in this study. Thus, those persons with an annual disposable family income below $56 and above $85,641 have been excluded from the analyses. The distribution of the 98%-tile cohort shown in plot C and disposable family income plotted by the corresponding amount of family assets is shown in plot D.

All analyses conducted in this report are based on the 98%-tile unless otherwise stated.

#### Covariates

##### Parental educational attainment

Information on parental educational attainment was delineated at the year of the cohort members's 1$^{st}$ birthday. It was defined at the maximum of maternal and paternal educational attainment. It was categorised into three groups:

- **Low**: Mandatory school, corresponding to a maximum length of education of seven years for persons born before January 1, 1958, and nine years for persons born on or after that date
- **Middle**: Secondary school and vocational education and short-cycle higher education, which approximated to a maximum of 10-12 years of schooling
- **Higher**: Medium- or long cycle higher education, approximately over 12 years of education

##### Family cohabitation status

Family's cohabitant status in the year before cohort member's 15$^{th}$ birthday was defined and categorized as

1. Cohabiting or married
2. Single
3. Single, living alone

##### Origin

Defined as parents' country of birth

1. Both parents born in Denmark
2. One or both parents born abroad

##### Urbanicity

Measure for urbanization was delineated using population density.

1. Capital
2. Capital suburb
3. Provincial city
4. Provincial towns
5. Rural area

##### Parental psychiatric history

Defined as either parent having received a psychiatric diagnosis in the Psychiatric Central Research Register (ICD-10 F-chapter [F00-F99] and equivalent ICD-8 codes [290-315]) before cohort member's 15$^{th}$ birthday.

##### Parental incarceration

Defined as either parent being convicted with a custodial sentence between cohort member's birth and 15$^{th}$ birthday.

#### Creation of simulated cohorts

To explore whether there is enough variability in disposable family income between siblings within the same family, we created a simulated cohort. Here, we group individuals into families at random. First we considered the 404 179 person (62.8% of the cohort) across 185 558 families (43.6% of the families) who had a least one full sibling in the full cohort (and were thus included in the sibling comparison design). For each of the 185 558 families, we picked one the siblings at random (n = 185 558) and each remaining sibling (n = 218 620) was replaced by a random individual (from the sibling cohort of n = 404 179) who had a similar family disposable income, the same sex, birth year, and birth order, and who came from a family with the same number of siblings. The difference in disposable income between the original sibling and the replaced random one was below $100 for 213 315 (97.6%) of the individuals, ($100-$200 for 2 783 (1.3%) of the individuals), $200-$500 for 1 611(0.7%), and above $500 for 911 (0.04%) of the individuals. We created a simulated cohort where one sibling of each of the 186 588 families was linked to random individuals with similar characteristics of the real sibling, but who were not family members.To examine whether the regular cohort and the sibling comparison design might be estimating different parameters, we also created two additional simulated cohort. The first in which the replaced siblings were similar to the real ones in all parameters (sex, birth year, birth order and number of sibling) but were not restricted to have a similar income. In the second the pseudo-sibling were chosen completely at random. In these two simulated cohorts, one would expect to find similar estimates in the entire cohort and in the sibling comparison design. The results are shown in the below table.

**Table. Adjusted hazard ratios (aHR) and 95% confidence intervals (CI) of being diagnosed with severe mental disorders for each $15 000 increase in disposable family income for different criteria for pseudo-sibling selection.**

| Sibling population with simulated families | Full cohort analysis aHR (95%CI) | Sibling comparison analysis aHR (95%CI) |
|---|---|---|
| Pseudo-siblings chosen completely at random | 0.76 (0.73-0.79) | 0.76 (0.72-0.80) |
| Pseudo-siblings with same characteristics on sex, birth year, birth order and family size as original sibling | 0.78 (0.75-0.80) | 0.76 (0.72-0.80) |
| Pseudo-siblings with same characteristics on sex, birth year, birth order and family size and ***income*** as original sibling | 0.77 (0.74-0.80) | 0.93 (0.85-1.01) |

### Figures

***Figure S1*. Distribution of annual disposable family income. Panels A and B show the density distribution and disposable income by the amount of family assets for the full cohort. Panels C and D show the same measures after excluding those below the first percentile and those above the 99th percentile**

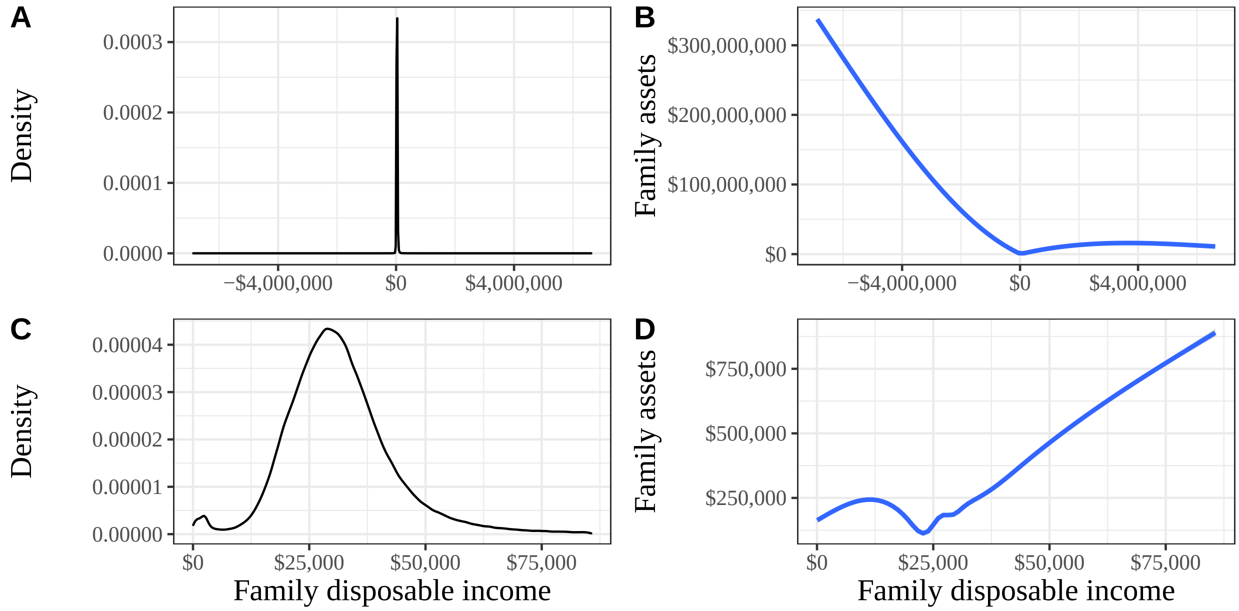

#### Figures

***Figure S2***. Illustration of three life course models

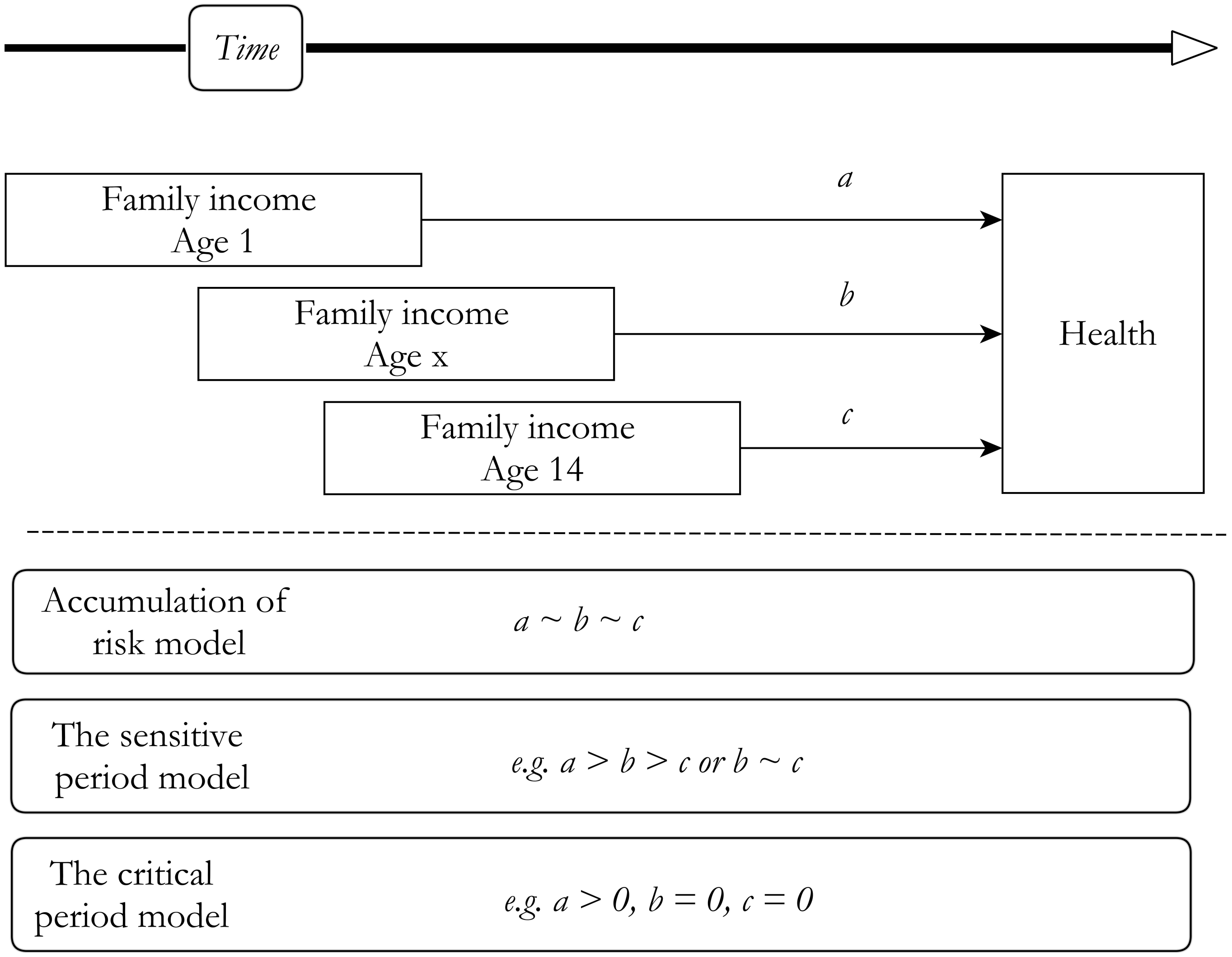

***Figure S3*. Adjusted hazard ratios (aHR) and 95% confidence intervals (CI) of being diagnosed with severe mental disorders for each $15 000 increase in disposable family income, grouping disposable family income as quintiles and deciles, respectively.**

| **Model** | **HR (95%CI)** |
|---|---|
| **Family income quintiles** | |
| Lowest quintile | Ref (−) |
| 2 | 0.86 (0.82−0.91) |
| 3 | 0.72 (0.68−0.76) |
| 4 | 0.66 (0.63−0.71) |
| Highest quintile | 0.62 (0.58−0.66) |
| **Family income deciles** | |
| Lowest decile | Ref (−) |
| 2 | 0.94 (0.88−1.00) |
| 3 | 0.83 (0.78−0.90) |
| 4 | 0.73 (0.68−0.79) |
| 5 | 0.69 (0.64−0.74) |
| 6 | 0.65 (0.60−0.70) |
| 7 | 0.62 (0.57−0.67) |
| 8 | 0.58 (0.53−0.62) |
| 9 | 0.56 (0.52−0.61) |
| Highest decile | 0.49 (0.45−0.54) |

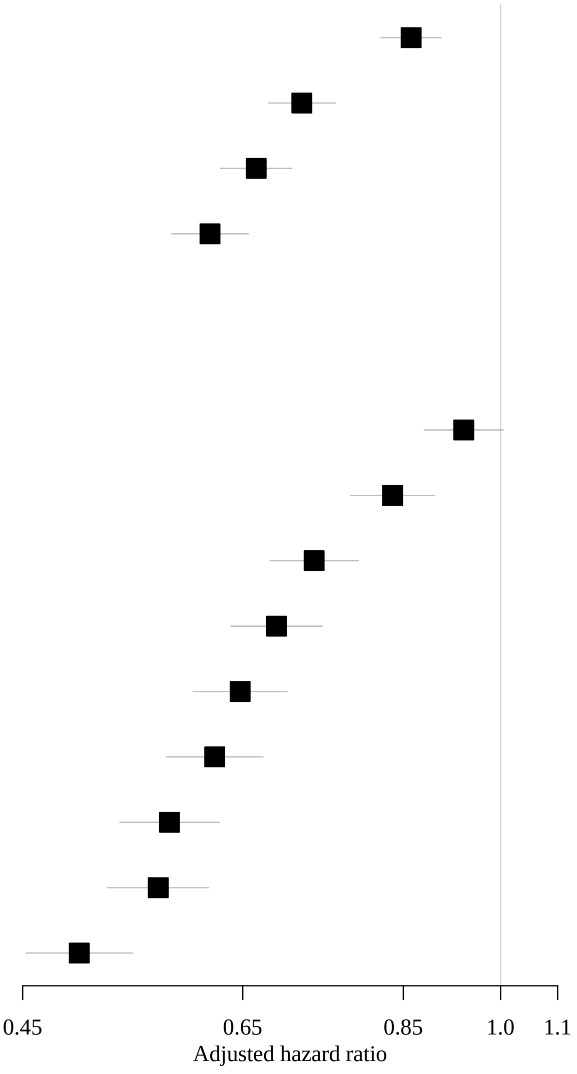

Note: The *Adjusted model* is adjusted for sex, birth order, birth year, origin, parental incarceration, parental psychiatric history, parental educational attainment, urbanicity and family cohabitation status.

***Figure S4*. Adjusted hazard ratios (aHR) and 95% confidence intervals (CI) of being diagnosed with severe mental disorders for each $15 000 increase in disposable family income, based on the entire cohort and the sibling cohort, respectively**

| Model | HR (95%CI) |
|---|---|
| Entire cohort | 0.78 (0.76−0.81) |
| Sibling cohort | 0.77 (0.74−0.80) |

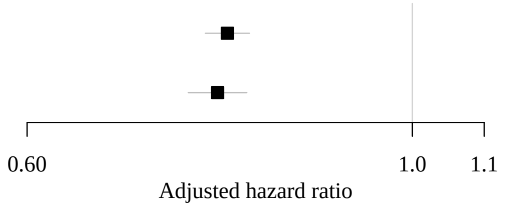

Note: The *Adjusted model* is adjusted for sex, birth order, birth year, origin, parental incarceration, parental psychiatric history, parental educational attainment, urbanicity and family cohabitation status.

***Figure S5*. Distribution of age difference between siblings in years with mean and interquartile range (IQR) ($n$ = 404,177)**

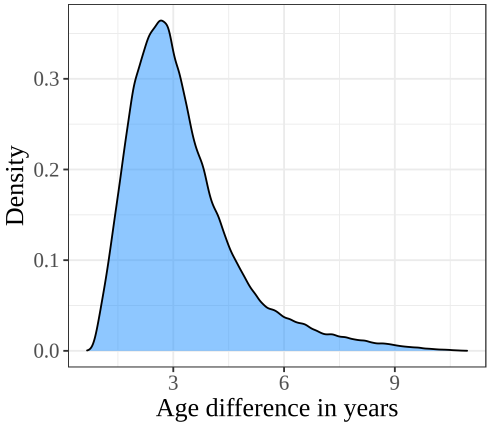

| **Mean (IQR)** |
|---|
| 3.2 (2.2 : 3.9) |

***Figure S6.*** **Annual disposable family income distribution with indication of income decile groupings with mean, median and interquartile range (IQR) ($n$ = 404,177)**

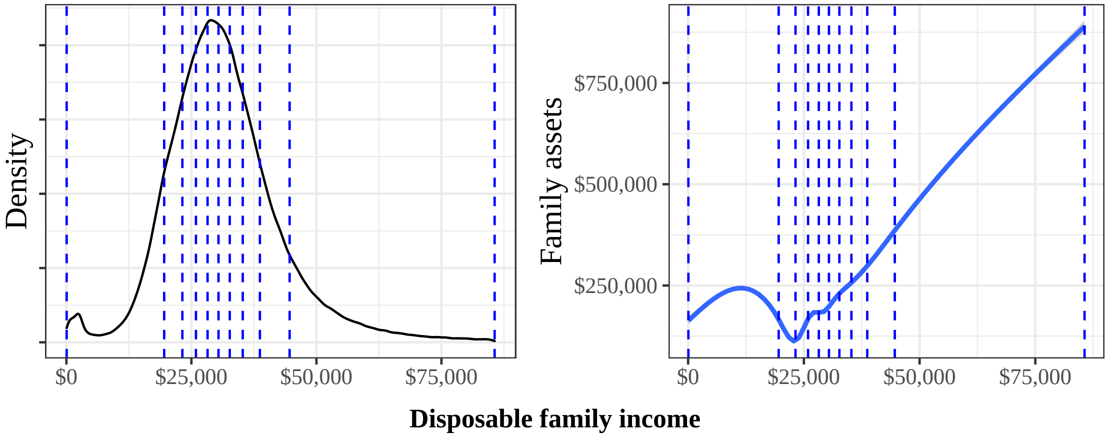

| Mean (IQR) |
|---|
| $30,712 ($24,310 : $35,741) |

***Figure S7.*** **Overall (left) and absolute (right) annual disposable family income differences between siblings measured at age 14 with mean, median and interquartile range (IQR) ($n$ = 404,177)**

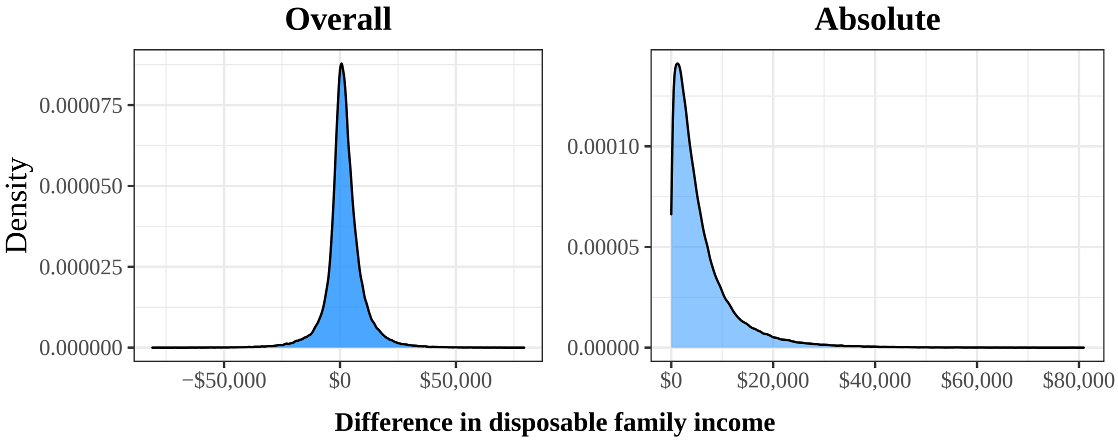

| | Overall | Absolute |
|---|---|---|
| Mean | $1,873 | $5,957 |
| Median | $1,495 | $3,945 |
| IQR | -$1544 : $5,211 | $1,807 : $7,701 |

***Figure S8*. Disposable family income distribution comparing the sibling cohort according to parental educational levels**

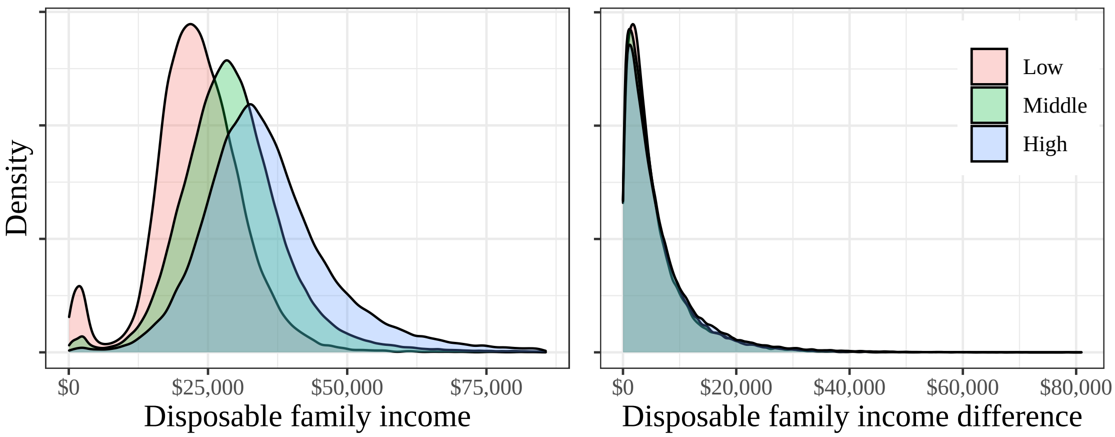

| | Family income for parental education levels | | | Difference between siblings for parental education levels | | |
|---|---|---|---|---|---|---|
| | **Low** | **Middle** | **High** | **Low** | **Middle** | **High** |
| Mean | $23,267 | $29,451 | $35,584 | $5,565 | $5,798 | $6,301 |
| Median | $23,045 | $28,827 | $34,089 | $3,703 | $3,892 | $4,131 |
| IQR | $18,597 : $28,021 | $23,607 : $34,397 | $28,100 : $41,295 | $1,801 : $7,175 | $1,788 : $7,511 | $1,846 : $8,169 |

***Figure S9*. Disposable family income distribution comparing the sibling cohort with and without a first-born child diagnosed with severe mental disorders**

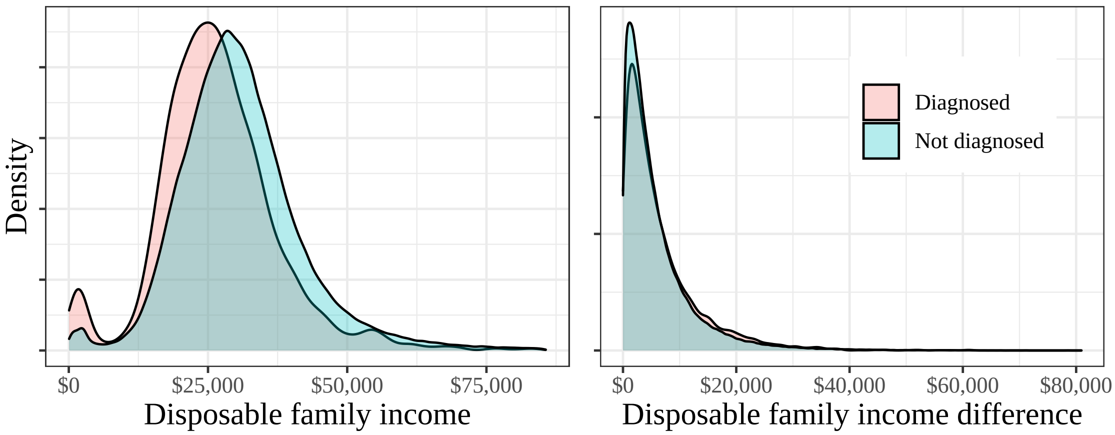

| | Family income | | Difference between siblings | |
|---|---|---|---|---|
| | **Diagnosed** | **Not diagnosed** | **Diagnosed** | **Not diagnosed** |
| Mean | $26,675 | $30,757 | $6,444 | $5,951 |
| Median | $25,897 | $29,732 | $4,224 | $3,943 |
| IQR | $20,329 : $32,107 | $23,830 : $36,246 | $1,824 : $8,598 | $1,806 : $7,692 |

### Tables

##### ***Table S1*. Overview over the three cohorts used in this study**

| | **Original cohort** | **Original cohort** | **Simulated cohort** |
|---|---|---|---|
| | Full cohort | Sibling cohort | Pseudo-sibling cohort |
| Cohort members | All individuals born in Denmark between 1986-1996 | Individuals with at least one sibling in the full cohort | One sibling from each family from the sibling cohort (chosen at random). Other sibling were randomly replaced by other individuals from the sibling cohort with similar characteristics |
| N (individuals) | 643 814 | 404 179 | 404 179 |
| N (families) | 425 193 | 185 558 | 185 558 |
| Standard Cox analysis | Model 1[a] | Model 5[a] | Model 3[a] |
| Sibling comparison design | - | Model 2 | Model 4 |

[a] T*he Standard Cox analyses* are Cox proportional hazard models with age as the underlying time scale and adjusted for sex, birth order, birth year, origin, parental incarceration, parental psychiatric history, parental educational attainment, urbanicity and family cohabitation status.

[b] *The Sibling comparison designs* are the standard Cox analyses described above but with the baseline hazard stratified by family.

***Table S2.*** **Person-time at risk, number of cases and rates per 1000 person-years among individuals born in Denmark 1986-1996 and followed up until 31 December 2018**

| | **Number of diagnoses** | **Person-time at risk (years)** | **Rate per 1000 person-years** |
|---|---|---|---|
| Overall | 14,637 | 7,787,305 | 1.88 |
| **Disposable family income decile** | | | |
| Lowest decile | 2,560 | 775,055 | 3.30 |
| 2 | 2,116 | 786,620 | 2.69 |
| 3 | 1,760 | 790,372 | 2.23 |
| 4 | 1,461 | 793,067 | 1.84 |
| 5 | 1,323 | 792,339 | 1.67 |
| 6 | 1,211 | 790,724 | 1.53 |
| 7 | 1,159 | 782,903 | 1.48 |
| 8 | 1,055 | 771,973 | 1.37 |
| 9 | 1,054 | 758,926 | 1.39 |
| Highest decile | 938 | 745,325 | 1.26 |

***Table S3*. Comparison between individuals with and without siblings**

| | | | Disposable family income | |
|---|---|---|---|---|
| | **n** | **Severe mental disorder(%)** | **Mean** | **Median (IQR)** |
| No | 239 635 | 5 721 (2.4%) | $32,766 | $31,594 ($25,248 : $38,730) |
| Yes | 404 179 | 8 916 (2.2%) | $30,712 | $29,816 ($24,310 : $35,741) |

***Table S4.*** **Comparison of characteristics of families with large versus small differences in disposable family income between siblings at age 14**

| Variable | $0 - $5,000 | $5,001 - $10,000 | $10,001 - $15,000 | $15,000 - $30,000 | $30,001 - $45,000 | Above $45,000 |
|---|---|---|---|---|---|---|
| Overall | 238 924 (59.1%) | 96 579 (23.9%) | 35 962 (8.9%) | 27 728 (6.9%) | 4 112 (1%) | 874 (0.2%) |
| **Parental Educational level** | | | | | | |
| Low | 19282 (8.1%) | 7158 (7.4%) | 2521 (7%) | 2070 (7.5%) | 169 (4.1%) | 30 (3.4%) |
| Middle | 128453 (53.8%) | 51771 (53.6%) | 18677 (51.9%) | 13454 (48.5%) | 1978 (48.1%) | 422 (48.3%) |
| High | 76756 (32.1%) | 32278 (33.4%) | 12830 (35.7%) | 10437 (37.6%) | 1699 (41.3%) | 372 (42.6%) |
| Missing | 14433 (6%) | 5372 (5.6%) | 1934 (5.4%) | 1767 (6.4%) | 266 (6.5%) | 50 (5.7%) |
| **Disposable family income decile** | | | | | | |
| Lowest decile | 25654 (10.7%) | 7813 (8.1%) | 3313 (9.2%) | 4897 (17.7%) | 391 (9.5%) | 0 (-) |
| 2 | 27959 (11.7%) | 9891 (10.2%) | 2767 (7.7%) | 1561 (5.6%) | 329 (8%) | 0 (-) |
| 3 | 27921 (11.7%) | 10400 (10.8%) | 3051 (8.5%) | 1464 (5.3%) | 157 (3.8%) | 36 (4.1%) |
| 4 | 28122 (11.8%) | 10439 (10.8%) | 3204 (8.9%) | 1490 (5.4%) | 108 (2.6%) | 42 (4.8%) |
| 5 | 26985 (11.3%) | 10713 (11.1%) | 3318 (9.2%) | 1606 (5.8%) | 149 (3.6%) | 39 (4.5%) |
| 6 | 26063 (10.9%) | 10494 (10.9%) | 3513 (9.8%) | 1874 (6.8%) | 160 (3.9%) | 35 (4%) |
| 7 | 24707 (10.3%) | 9936 (10.3%) | 3744 (10.4%) | 2198 (7.9%) | 166 (4%) | 41 (4.7%) |
| 8 | 21655 (9.1%) | 9837 (10.2%) | 4076 (11.3%) | 2845 (10.3%) | 346 (8.4%) | 61 (7%) |
| 9 | 18130 (7.6%) | 9249 (9.6%) | 4201 (11.7%) | 3691 (13.3%) | 675 (16.4%) | 126 (14.4%) |
| Highest decile | 11728 (4.9%) | 7807 (8.1%) | 4775 (13.3%) | 6102 (22%) | 1631 (39.7%) | 494 (56.5%) |
